# Arsenic in drinking-water sources in Bangladesh, 2012-13 to 2025: national trends, geographic persistence and implications for well testing

**DOI:** 10.64898/2026.10.06.26364882

**Authors:** S. M. Ashraf Ul Hoque, Md Khairul Islam, Mohammad Khalid Hasan, Mohiuddin Sharif, Tanzin Naher, Bilkis Banu, Zenat Zebin Hossain, Kamran ul Baset

## Abstract

**Background:** Arsenic in tube-well water remains a major health hazard in Bangladesh. National surveys suggest falling contamination, but changing field kits and small district samples leave the size and geography of the fall uncertain. We aimed to estimate how arsenic contamination of household drinking-water sources in Bangladesh changed between 2012-13 and 2025, nationally and across districts.

**Methods:** We analysed 11,624 household source-water tests from three nationally representative Multiple Indicator Cluster Surveys in 2012-13, 2019 and 2025. The main outcome was a field-kit reading above 10 parts per billion (ppb); a reading above 50 ppb was secondary. We adjusted the 2012-13 readings with that survey’s laboratory cross-check. Design-based models estimated prevalence change and adjusted prevalence ratios, testing sensitivity to later-kit error and interview month. We assessed whether districts high in both earlier rounds still held people exposed in 2025.

**Results:** Household prevalence above 10 ppb fell from a laboratory-adjusted 24.0% in 2012-13 to 14.4% in 2025, a fall of 9.6 percentage points (95% CI 7.7 to 11.4) and an adjusted prevalence ratio of 0.61 (0.55 to 0.67). Changes above 50 ppb and since 2019 were uncertain after adjustment for interview month. About 26 million people (24 to 28 million) still lived in households whose source exceeded 10 ppb. The 25 districts high in both earlier rounds held 38.6% of the population but 71.9% (68.0 to 75.8) of these people.

**Conclusions:** Arsenic contamination of drinking-water sources in Bangladesh appears to have fallen since 2012-13. However, the size of this fall is uncertain, because the field kits used in 2019 and 2025 were never checked against laboratory measurements. Exposure remained concentrated in the districts that had high prevalence in earlier surveys. Well testing could start in these districts, while testing continues in the rest of the country. Future surveys need laboratory-validated kits.

## Introduction

Arsenic contamination of groundwater in Bangladesh has been called “the largest mass poisoning of a population in history” [1]. Tube wells installed to provide microbiologically safe water were found in 1993 to draw naturally arsenic-rich water from shallow aquifers [1,2]. The provisional World Health Organization (WHO) guideline value for arsenic in drinking water is 10 micrograms per litre (µg/L), equal to 10 parts per billion (ppb) [3]. However, the national standard in Bangladesh remains 50 ppb [4].

Chronic arsenic exposure causes cancers of the skin, lung and bladder and contributes to cardiovascular, neurological and developmental disease [5]. The Health Effects of Arsenic Longitudinal Study followed adults in Araihazar, near Dhaka. There, the risk of premalignant skin lesions was already raised at 8.1 to 40 µg/L, a range that includes the WHO guideline [6]. The same cohort showed higher cardiovascular and all-cause mortality with higher exposure [7,8]. Drinking water above 50 µg/L during pregnancy was associated with a 14% higher risk of fetal loss and a 17% higher risk of infant death [9]. Using the 2009 national survey, Flanagan and colleagues attributed about 24,000 adult deaths a year to arsenic above 50 µg/L [10].

National estimates fell from 57 million people above 10 µg/L in 2000 to 40 million in 2013 [11]. Government retesting of wells in 2021-23 suggested that about 20 million people still drank unsafe water [12]. Contamination has also long been uneven across the country, and a Bayesian model of the 2012-13 samples found high risk in the northeast and southwest [13]. Under the

Sustainable Development Goals, safely managed drinking water must be free from priority chemical contamination, including arsenic, judged against national standards [14].

The Bangladesh Multiple Indicator Cluster Survey (MICS), a national household survey, tested drinking-water sources for arsenic in 2012-13, 2019 and 2025 [15–17]. It can therefore show whether contamination kept falling, and where. Two features of MICS data make these questions hard to answer. First, the field kits vary between rounds, and the accuracy of kits is a concern [18], so a perceived drop could reflect the kits. Second, district samples are small, with approximately 40 to 50 sampled clusters per district [15–17]. As a result, a district in the top tier in one round may not differ reliably from districts ranked just below it. Policy makers therefore lack a comparable 2025 figure and do not know whether earlier high districts remain the right places to start testing [10,12].

We aimed to estimate how arsenic contamination of household drinking-water sources in Bangladesh changed between 2012-13 and 2025, nationally and across districts. To make the rounds comparable, we adjusted the 2012-13 readings with that survey’s laboratory table and tested how much error in the 2019 and 2025 kits would remove the change.

## Methods

### Study design and data

This secondary analysis used three repeated, nationally representative cross-sectional household surveys: MICS5 (2012-13), MICS6 (2019) and MICS7 (2025) [15–17]. The 2025 round also recorded tube-well depth and cluster coordinates, displaced within the upazila (sub-district) for privacy [17]. We followed the Strengthening the Reporting of Observational Studies in Epidemiology (STROBE) guideline [19].

### Sampling and arsenic testing

Each survey used a stratified two-stage cluster design, selecting census enumeration areas with probability proportional to size and then 20 households in each. Fieldwork ran from December to April in 2012-13, January to June in 2019 and February to June in 2025 (S1 Text). A random subsample of households had their main drinking-water source tested: one per cluster in 2012-13 and 2019, and two in 2025 (S1 Text). Trained measurers tested each sample in the field with a kit, and blank tests, which should read zero, checked quality (S1 Table). Each result was recorded with the household interview (S1 Text). We analysed the 11,624 households with a valid result (S1 Fig), none of which lacked a model covariate.

### Outcomes and the field kits

Following the MICS 2025 definitions [17], the main outcome was a reading strictly above 10 ppb, based on the WHO guideline [3]. A reading strictly above 50 ppb, the national standard [4], was a secondary outcome. Each kit gives readings on a fixed scale of values. In 2012-13 measurers used the Arsenic Econo-Quick kit and took the higher value when a colour fell between chart colours [15]. Because the 2012-13 scale stepped from 10 to 25 ppb, any reading above 10 ppb was 25 ppb or more. The reports do not name the later kits or their reading rules (S1 Text). As a sensitivity analysis, we counted later readings of exactly 10 (or 50) ppb as exceeding (S1 Table).

The 2012-13 report cross-checked kit readings against laboratory analysis of stored quality-control samples. For each reading it published the share of samples in four laboratory ranges (S2 Table) [15]. We used this table to give each 2012-13 household a probability that its source truly exceeded each cut. For example, 36% of samples read as 25 ppb were above 10 ppb in the laboratory, so such a household had a probability of 0.36. The weighted mean of these probabilities is the laboratory-adjusted prevalence. This assumes that the table applies equally across source types, districts and covariates.

The 2019 and 2025 reports describe no laboratory check [16,17], so we used their readings as recorded. We tested how kit error would change the results (S1 Text, S3 Table). Assuming no false positives, we computed the later-kit sensitivity that would remove the fall since 2012-13. We also recomputed the ratios over a grid of later-kit sensitivity and specificity, and varied the laboratory table cells (S1 Text).

### Covariates

Covariates were urban or rural residence, wealth quintile, division (2025 boundaries) and interview month. Wealth quintiles came from the MICS wealth index of each round. The index is built by principal component analysis of household assets, dwelling characteristics, and water and sanitation facilities. Separate urban and rural scores are combined into one national score [16,17]. Tube-well users were households whose main source was a tube well; in 2025 deep and shallow tube wells were separate response codes.

### Statistical analysis

The primary analysis was the national change above 10 ppb from 2012-13 to 2025. The 50 ppb, 2019 to 2025 and sensitivity analyses test its robustness. District analyses show where change occurred, and the 2025 spatial and well-depth analyses help explain it. No protocol was registered, and several analyses were chosen during analysis (S1 Text). All estimates used the strata, clusters and water quality weights of each round (S1 Text). Household-weighted shares are the main estimates, because arsenic describes the source; member-weighted shares give the share of people.

For the national change, prevalence ratios came from survey-weighted quasi-Poisson regression with a log link [20], which accepts the fractional laboratory-adjusted outcome. Models adjusted for urban residence, wealth quintile and division, so the adjusted ratios are associations with survey round, not causal effects. Because changes in population make-up may be part of the real trend, our main estimate is the percentage-point change in national prevalence. We separated this change into within-source and source-mix components (S4 Table). In sensitivity analyses, we added interview month in five groups, or kept only February to April, the months present in every round.

For exposed people, we counted those living in households whose tested source exceeded a threshold; this categorises the source, not individual intake or cumulative dose. We multiplied the member-weighted 2025 prevalence and its bounds by the 2022 census population of 169.8 million, adjusted for net coverage error [21]. The counts therefore apply 2025 prevalence to the 2022 population.

For the district analysis, we kept 62 of 64 districts with at least 30 tests in all rounds; Narayanganj and Rangamati were left out. A district was high in a round if its kit-reading prevalence above 10 ppb was at or above the median of the 62. This is an operational class and not a health threshold. District intervals used the Korn and Graubard method [22]. Our main targeting set was the districts high in both the 2012-13 and 2019 rounds. It makes no use of the 2025 data, so scoring it on 2025 is a retrospective temporal evaluation of a rule chosen during analysis. Survey ratio estimators gave each set’s share of the 2025 population and of exposed people. A set built from the 2009 national drinking water quality survey [23] and other robustness checks are described in S1 Text and S5 Table.

To see whether high districts formed local clusters, we used the Getis-Ord Gi* statistic [24] on geoBoundaries district boundaries [25]. We used permutation P values and Benjamini-Hochberg false discovery rate control [26]. We also computed global Moran’s I [27] of district prevalence. For 2025, we aggregated tests to 3,138 sampled clusters, unweighted, so maps describe sampled clusters, not population prevalence. Spatial clustering was Moran’s I of residuals from binomial models. The models added urban residence, wealth, elevation and rainfall, then division, and then a thin plate regression spline of the coordinates in place of division [28]. Among tube-well households, survey-weighted quasi-Poisson models compared deep with shallow wells, adjusted for urban residence, wealth and division, and then with a depth by division interaction.

We performed all analyses using Stata version 17 (StataCorp LLC, College Station, TX, USA) and R version 4.6.1 (R Foundation for Statistical Computing, Vienna, Austria).

### Ethics

We used anonymised microdata from the United Nations Children’s Fund (UNICEF) MICS programme, accessed on 26 September 2026 under the MICS data-use terms. The authors had no access to information that could identify individual participants during or after data collection.

The International Centre for Diarrhoeal Disease Research, Bangladesh (icddr,b) Ethical Review Committee approved the 2025 protocol in November 2024 (reference PR-24113). A government technical committee led by the Bangladesh Bureau of Statistics approved the 2019 protocol.

Respondents gave verbal consent. The 2012-13 report documents no ethics review. No further ethical approval was required for this analysis. We used coordinates only at cluster level.

## Results

### Study population

Valid results were obtained for 2,554 of 2,760 planned tests in 2012-13 (92.5%), 3,028 of 3,220 in 2019 (94.0%) and 6,042 of 6,298 in 2025 (95.9%; S1 Fig). Tested households became more urban (20.8% to 30.2%), and fewer used a tube well (91.4% to 71.8%; Table 1). Our estimates reproduced the published 2025 and 2019 tables exactly and the 2012-13 values within 0.6 points after laboratory adjustment (member-weighted; S6 Table). In 2019, paired source and drinking-water tests agreed at 10 ppb in 98% of households (S1 Text).

**Table 1.** Characteristics of households with a valid source-water arsenic test, by survey round (weighted percent).

| **Characteristic** | **2012-13** | **2019** | **2025** |
|---|---|---|---|
| Households tested, n | 2,554 | 3,028 | 6,042 |
| Urban residence | 20.8 | 22.1 | 30.2 |
| Main source is a tube well | 91.4 | 85.8 | 71.8 |
| Wealth quintile 1 (poorest) | 21.1 | 19.9 | 21.0 |
| Wealth quintile 2 | 20.9 | 20.8 | 19.9 |
| Wealth quintile 3 | 18.4 | 20.4 | 19.4 |
| Wealth quintile 4 | 20.0 | 20.4 | 19.8 |
| Wealth quintile 5 (richest) | 19.5 | 18.5 | 19.9 |

*Percentages use household weights, and n is unweighted. Wealth quintiles are relative within each round.*

### Change in national prevalence

On kit readings, the share of households above 10 ppb fell from 26.6% in 2012-13 to 16.8% in 2019 and 14.4% in 2025 (Figure 1). After laboratory adjustment, the 2012-13 share was 24.0% (95% confidence interval [CI] 22.6 to 25.5). The share above 10 ppb therefore fell by 9.6 percentage points (95% CI 7.7 to 11.4) by 2025. The adjusted ratio was 0.61 (0.55 to 0.67; Figure 2). Of the 9.6-point fall, 7.0 points arose within source types and 2.6 from the shift away from tube wells. Among tube-well users, the share above 10 ppb fell from 25.2% to 18.0% (S4 Table).

**Figure 1.**
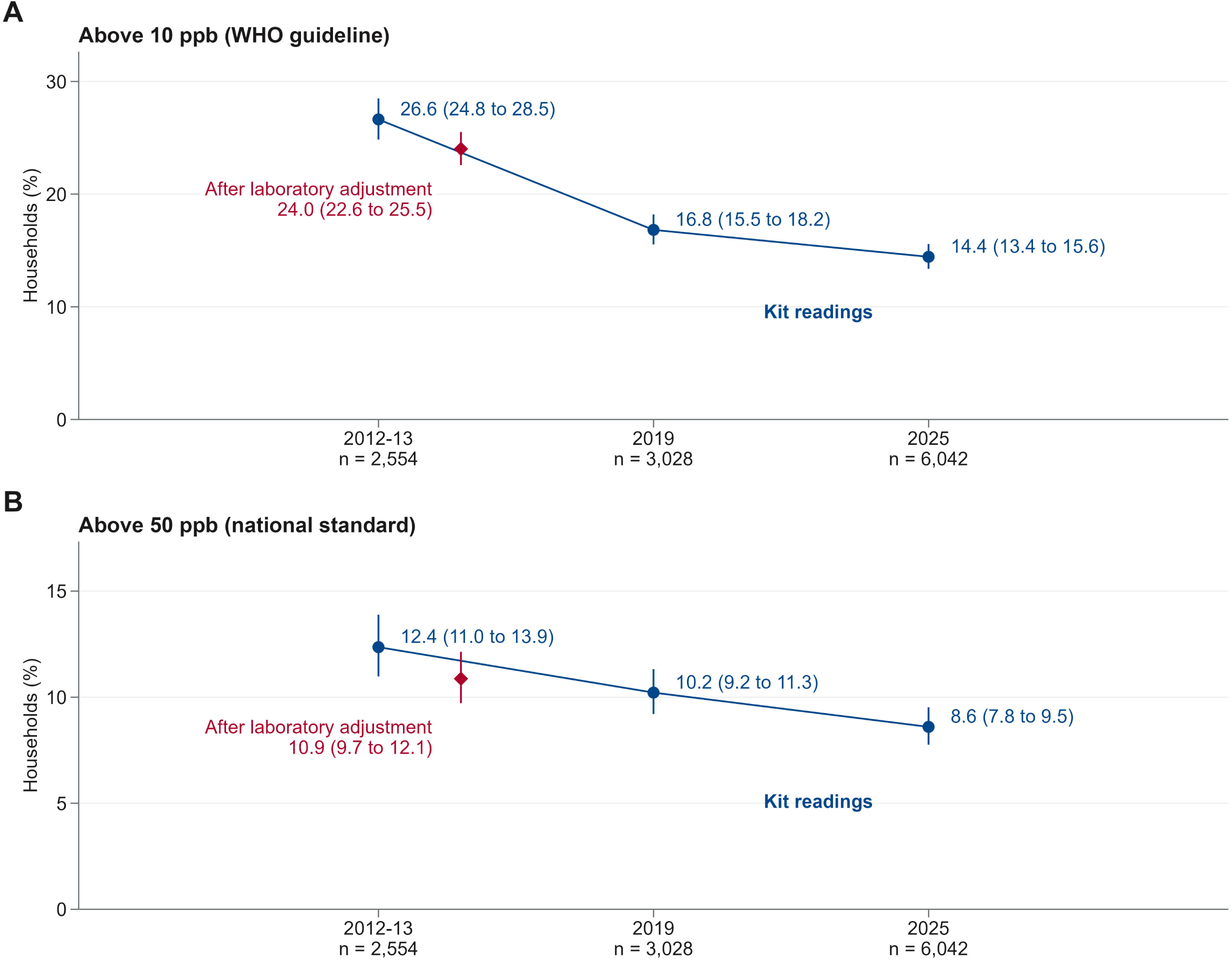
Arsenic in household drinking-water sources in Bangladesh, 2012-13 to 2025. Panel A shows the percentage of households whose source exceeded 10 ppb, and panel B the percentage above 50 ppb. Navy points are field-kit readings. Crimson diamonds show the 2012-13 value after adjustment with the survey’s laboratory cross-check, offset to the right for visibility. Vertical bars are 95% confidence intervals. They reflect survey sampling only and do not include error in the laboratory table.

**Figure 2.**
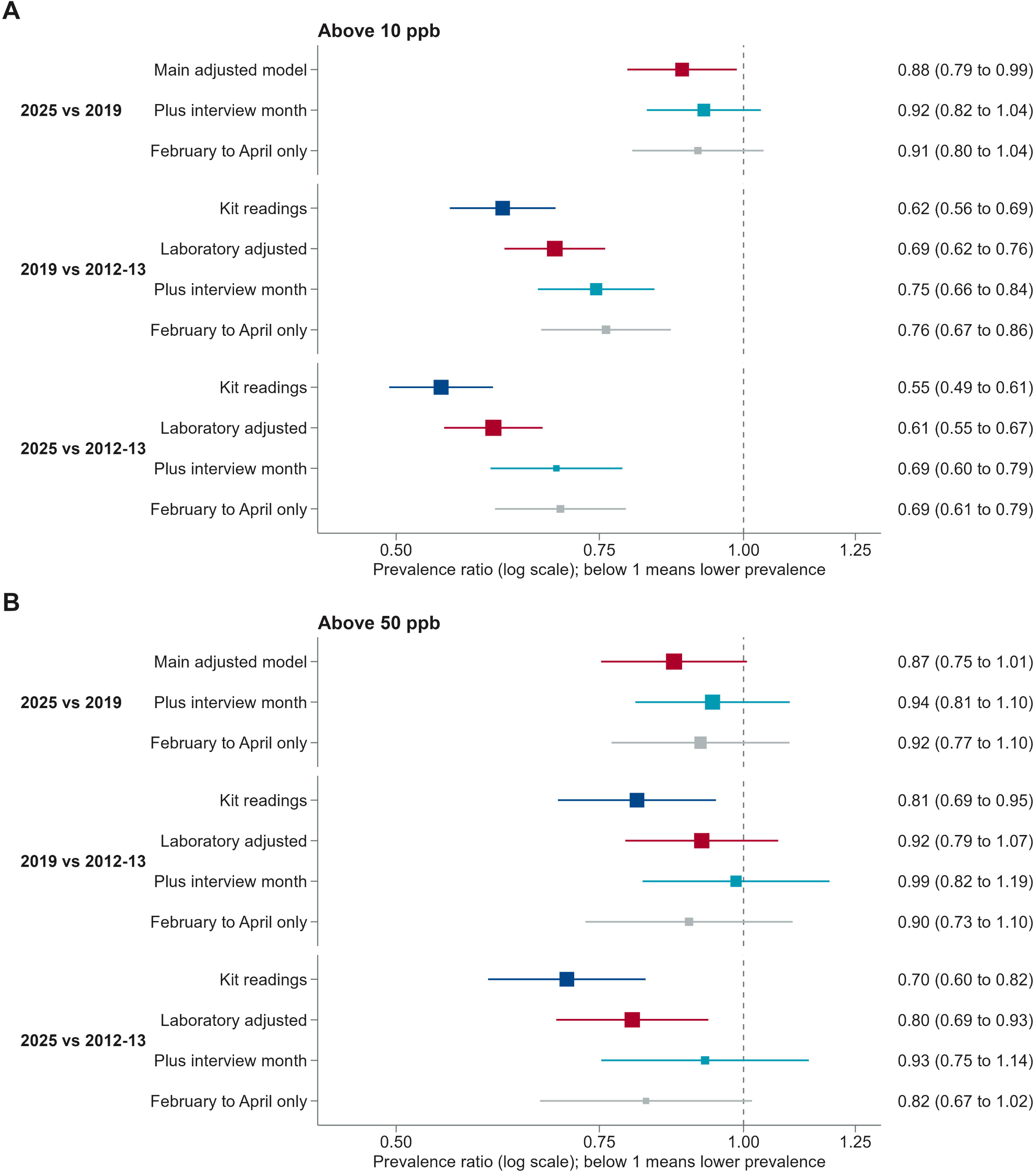
Adjusted prevalence ratios comparing survey rounds, for arsenic above 10 ppb. **(A)** and above 50 ppb (B). Each square is a ratio with its 95% confidence interval, and larger squares are more precise. All models adjust for urban residence, wealth quintile and division. Navy squares use field-kit readings in every round. Crimson squares replace the 2012-13 readings with their laboratory-adjusted values. The 2019 and 2025 readings had no laboratory adjustment, so for 2025 against 2019 the crimson square is the main adjusted model on kit readings. Teal squares add interview month (five groups) to the crimson model. Grey squares restrict the crimson model to interviews in February to April (1,283, 2,155 and 3,672 households). Intervals show survey sampling error only. The dashed line marks a ratio of 1, meaning no change.

Above 50 ppb, the laboratory-adjusted share fell from 10.9% to 8.6%, by 2.3 points (0.8 to 3.8), with a ratio of 0.80 (0.69 to 0.93; Figure 1B and Figure 2). With interview month in the model, the ratios weakened to 0.69 (0.60 to 0.79) at 10 ppb and 0.93 (0.75 to 1.14) at 50 ppb (Figure 2).

Readings of exactly 10 ppb were far more common in 2012-13 (31.9% of tests, against 3.4% in 2019 and 2.6% in 2025; S1 Table). In the laboratory, 89% of 2012-13 readings of 10 ppb were truly at or below 10 ppb (S2 Table).

The size of the fall therefore depends on the later kits. Assume that the laboratory table is correct and that the later kits give no false positives. Then the fall at 10 ppb would vanish if they detected only 60% (55 to 66) of sources truly above 10 ppb. If the 2012-13 share was at its lowest laboratory-table value, 69% would be enough (S3 Table). At 50 ppb, missing 21% (8 to 32) of such sources would remove the fall. Changes to the laboratory table gave crude ratios of 0.51 to 0.69 at 10 ppb (S3 Table).

From 2019 to 2025 the ratio was 0.88 (0.79 to 0.99) above 10 ppb and 0.87 (0.75 to 1.01) above 50 ppb. With interview month they were 0.92 (0.82 to 1.04) and 0.94 (0.81 to 1.10; Figure 2). If the 2019 and 2025 kits had different error rates, the 10 ppb ratio ranged from 0.60 to 1.19 (S3 Table). The 2019 to 2025 change did not differ by division (interaction P = 0.53 at 10 ppb), but the change across all three rounds did (P < 0.001; S1 Text). In 2025 the share above 10 ppb ranged from 1.8% in Barishal to 29.9% in Sylhet (Figure 3).

**Figure 3.**
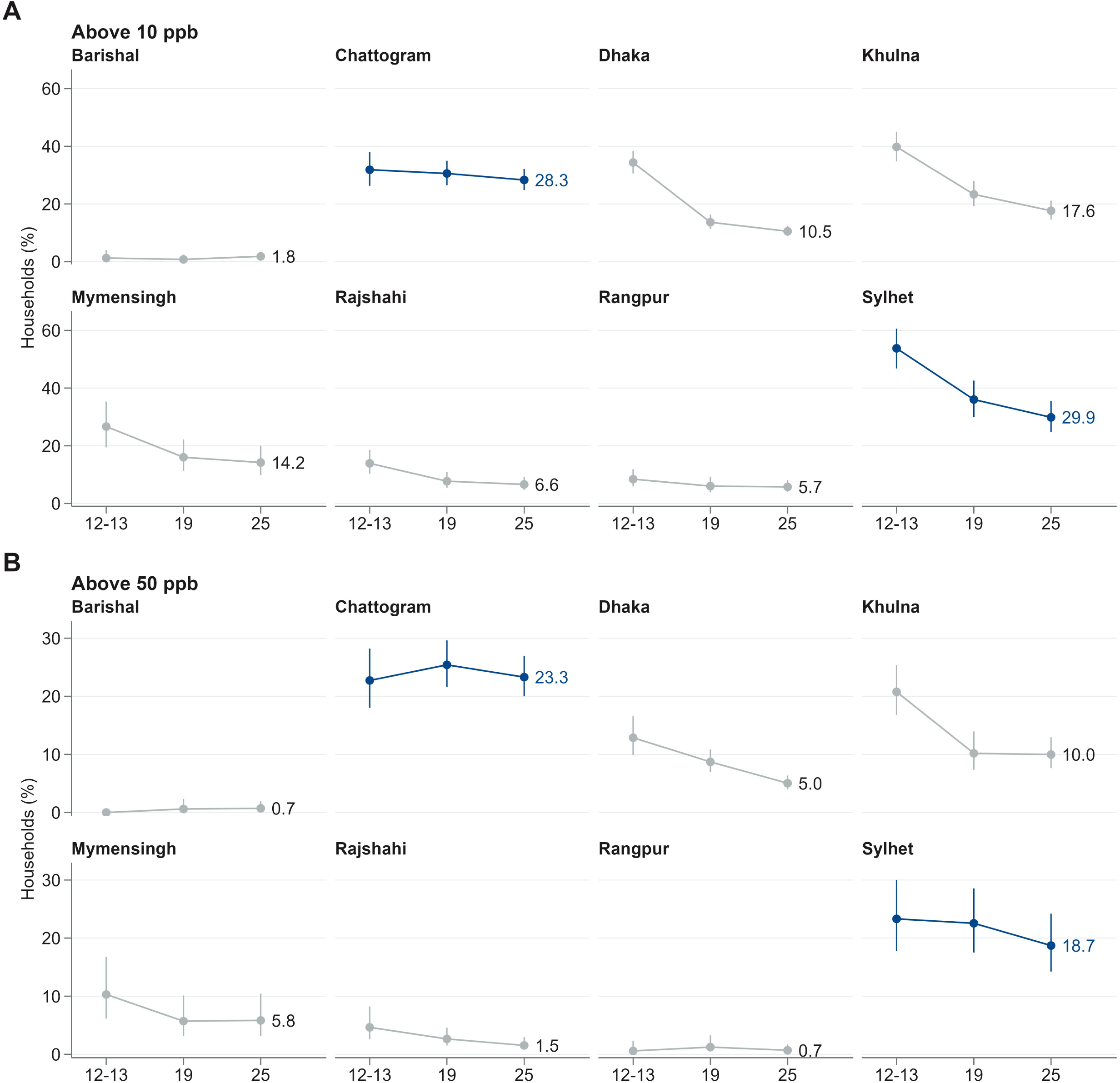
Arsenic in household drinking-water sources by division, 2012-13 to 2025. Panel A shows the percentage of households above 10 ppb and panel B above 50 ppb, with 95% confidence intervals where defined. Navy marks Chattogram and Sylhet, and the 2025 value is printed for each division. Barishal had no tests above 50 ppb in 2012-13, so no interval is shown. All values, including 2012-13, are household-weighted field-kit readings without laboratory adjustment, so the lines do not show validated trends.

### Change across districts and people exposed in 2025

In 2025, 15.1% (13.9 to 16.4) of people lived in households above 10 ppb (S6 Table). Applied to the 2022 census population, this means about 26 million people (24 to 28 million), or 23 to 32 million under illustrative kit-error scenarios (S3 Table). The district pattern changed little. Of the 62 districts, 23 were high in all three rounds and 24 in none (Figure 4A). Three 2012-13 district shares differed by more than 35 points from both the 2009 national survey and the 2019 round: Chandpur read 0.0%, Khagrachhari 84.1% and Tangail 82.3% (S7 Table). We flagged them as doubtful, and excluding them left the national crude ratio at 0.60 (0.54 to 0.66; S5 Table).

**Figure 4.**
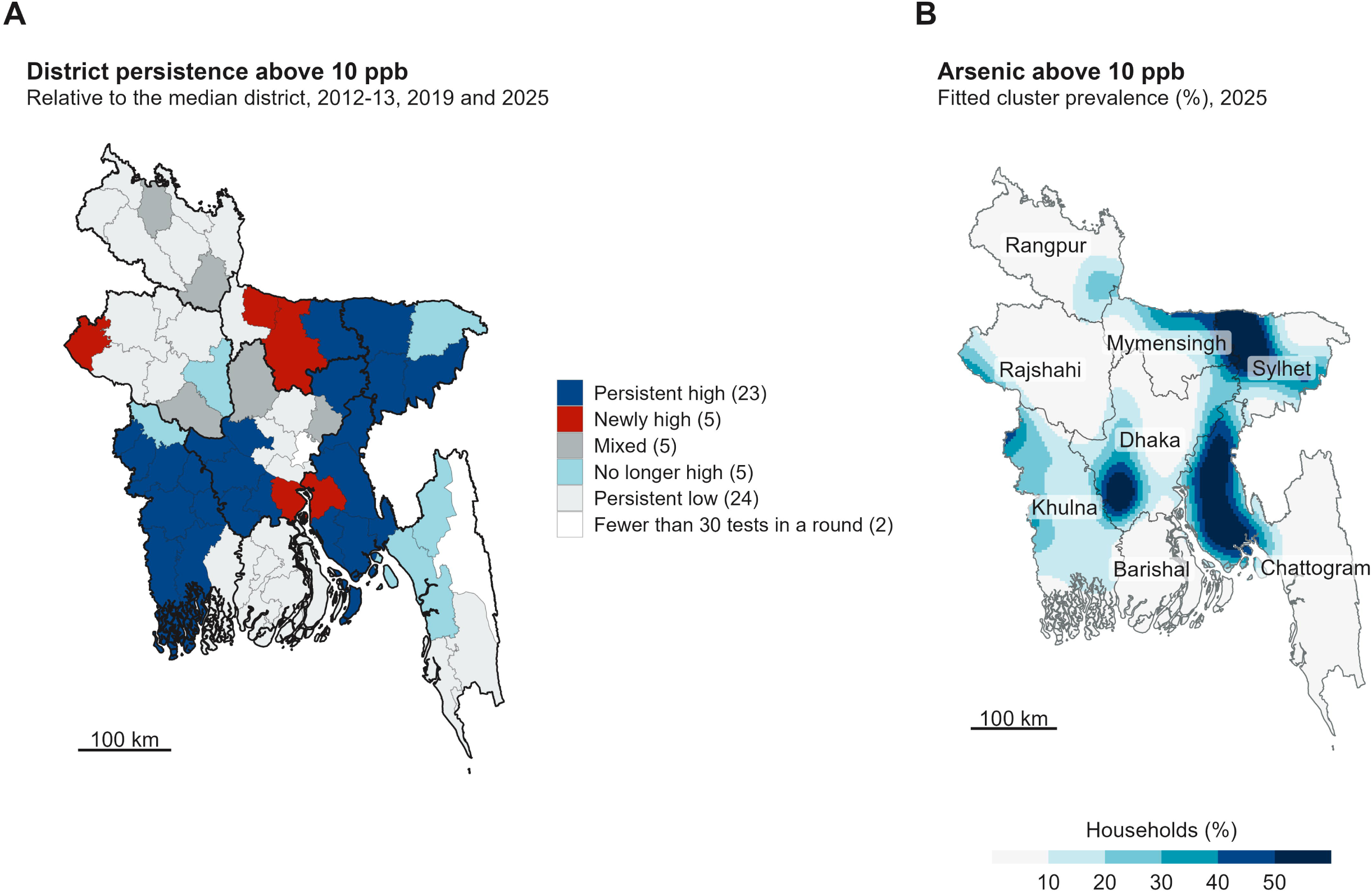
Where arsenic exposure persists. (A) District class across 2012-13, 2019 and 2025, relative to the median district in each round. Persistent high and persistent low districts were high, or low, in all three rounds. Newly high districts were not high in 2012-13 but high in 2025, no longer high districts showed the reverse, and mixed districts had other patterns. Districts with fewer than 30 tests in any round are left blank. Thick lines show division boundaries. (B) Fitted prevalence of arsenic above 10 ppb among sampled clusters in 2025, from a thin plate spatial smooth, in classes 10 percentage points wide. Division names are shown. Panel B describes sampled clusters at displaced locations and shows broad patterns only. It is not suitable for well-level targeting. Its sensitivity to the smoothing is shown in S3 Fig.

The 25 districts high in both 2012-13 and 2019 held 38.6% of the 2025 population but 71.9% (68.0 to 75.8, with the set fixed) of people above 10 ppb. They held 82.8% (78.7 to 87.0) of those above 50 ppb (Table 2). Re-deriving the set in 1,000 bootstrap samples gave 69.6% (59.8 to 75.9), which includes uncertainty in choosing the districts (S5 Table). The set was unchanged when 2012-13 districts were classed on laboratory-adjusted values (S1 Text). Replacing the class of the doubtful districts raised its share to 76.4% (72.6 to 80.2), and other definitions and a 2009-based set concentrated exposure similarly (S5 Table). In a descriptive comparison, exposed people outside the set were more often in the poorest quintile (28.4% against 17.6%; S5 Table).

**Table 2.** How well district classes from earlier rounds locate people exposed to arsenic in 2025.

| **District set** | **Districts** | **People in the set (%)** | **People above 10 ppb who live in the set (%, 95% CI)** | **People above 50 ppb who live in the set (%, 95% CI)** | **Above 10 ppb inside and outside the set (%)** | **Above 50 ppb inside and outside the set (%)** |
|---|---|---|---|---|---|---|
| Persistent high in all three rounds (uses 2025 data) | 23 | 31.3 | 68.9 (64.8 to 73.0) | 81.4 (77.0 to 85.7) | 33.2 and 6.8 | 23.8 and 2.5 |
| High in 2012-13 and 2019 (earlier rounds only) | 25 | 38.6 | 71.9 (68.0 to 75.8) | 82.8 (78.7 to 87.0) | 28.1 and 6.9 | 19.6 and 2.6 |
| High in 2019 (earlier round only) | 31 | 46.6 | 81.2 (77.6 to 84.9) | 92.2 (89.0 to 95.3) | 26.3 and 5.3 | 18.1 and 1.3 |

*Shares refer to the member-weighted 2025 population. A district is high in a round if its prevalence above 10 ppb is at or above the median. The median is taken over the 62 districts with at least 30 tests in every round. Shares are national, so the two unclassified districts (Narayanganj and Rangamati) count as outside each set. Intervals use the survey design and treat each set as fixed. The last two sets use no 2025 data. S5 Table gives bootstrap intervals that re-derive the sets. The 25 districts high in 2012-13 and 2019 are Brahmanbaria, Chattogram, Chuadanga, Cumilla, Faridpur, Feni, Gopalganj, Habiganj, Jashore, Jhenaidah, Khulna, Kishoreganj, Lakshmipur, Madaripur, Magura, Manikganj, Maulvibazar, Meherpur, Narail, Netrakona, Noakhali, Rajbari, Satkhira, Sunamganj and Sylhet.*

District ranks were stable between 2019 and 2025 (Spearman 0.90) but less so with 2012-13 (0.69 and 0.65; S1 Text). District shares in 2025 also matched those of the 2009 survey closely (Spearman 0.90 above 10 ppb; S8 Table). High districts lay near each other (global Moran’s I 0.35 to 0.40, all P < 0.001; S1 Text). Even so, no district was a local hotspot or coldspot after false discovery rate control (smallest q-value 0.12; S2 Fig).

### Spatial pattern and well depth in 2025

Among sampled clusters, arsenic was spatially clustered in 2025 (Moran’s I 0.349, P = 0.001). Urban residence, wealth, elevation and rainfall lowered this to 0.267, division to 0.169, and a spatial smooth in place of division to 0.049, which remained above chance (P = 0.001; S1 Text). Fitted prevalence was higher in the northeast around Sunamganj and Habiganj, and in a southeast belt from Cumilla to Noakhali (Figure 4B; S3 Fig). A smaller patch lay in the central south.

In 2025, 1,475 tube-well households reported a deep well and 3,198 a shallow well. Above 10 ppb were 12.4% (95% CI 10.5 to 14.5) of deep and 20.3% (18.6 to 22.1) of shallow wells. From a model without interaction, the adjusted ratio was 0.53 (0.44 to 0.63), and 0.42 (0.33 to 0.54) at 50 ppb. At 10 ppb the ratio differed by division (interaction P < 0.001; S9 Table). Deep wells had lower shares in Barishal, Chattogram, Dhaka and Khulna (ratios 0.07 to 0.57). Lower prevalence was not shown in the other four divisions, and in Mymensingh deep wells had a higher share (2.63, 1.36 to 5.12). Some division subgroups were small (S9 Table).

## Discussion

Arsenic contamination of household drinking-water sources in Bangladesh was lower in 2025 than in 2012-13, but the fall did not change where contamination was concentrated. The evidence was clearest at the WHO guideline, and weaker at the national standard and since 2019. Most of the 26 million people still exposed lived in districts that were already high.

The fall at 10 ppb survives the 2012-13 laboratory adjustment, but it is not proven. Earlier evaluations found field kits in Bangladesh erring in both directions [29–31]. In a controlled comparison, the Econo-Quick kit used in 2012-13 tended to underestimate near 10 µg/L [32]. Excess 2012-13 readings near 10 ppb more likely reflect how measurers read the colour chart, such as the rule of taking the higher value [15]. Later kits missing about two in five sources truly above 10 ppb, with no false positives, would erase the fall. These surveys cannot show whether such an error occurred. At 50 ppb a smaller error would be enough, so we cannot say whether the 50 ppb share fell.

The recent trend is less certain. Adding interview month moved the 2019 to 2025 ratio closer to 1, and in monitored wells arsenic varied little between seasons [33]. Month may therefore mainly reflect where teams worked and how they handled kits.

If the long-term trend is real, it may primarily reflect changes in the sources people use because monitored wells showed little change in arsenic levels over the years [33]. Declines were seen among tube-well users, and only about one-quarter of the total decline was due to the shift from tube wells. Other factors may include large-scale screening of nearly 5 million wells since 2000 and well-switching [34], as well as newly constructed wells [35].

Our estimate of about 26 million is lower than previous national estimates [11]. It is also about 12% to 28% of the 94 to 220 million people whom a global model predicted may drink groundwater above 10 µg/L [36]. That model used predicted concentrations and groundwater use, not tests, so the comparison is only approximate. It is higher than the 20 million from government retesting in 2021-23 [12], which used different methods and denominators. Even an uncertain fall may matter for health. In Araihazar, adults whose urinary arsenic fell below the baseline median had lower chronic disease mortality than those whose levels stayed high. The hazard ratio was 0.46 (95% CI 0.39 to 0.53) [37]. Those declines followed intensive community mitigation, which national programmes may not match.

Exposed people remained concentrated in high-risk districts, as reported previously in the 2009 national survey [23] and in a spatial analysis of the 2012-13 data [13]. High-risk districts remained high, yet no district was a local hotspot. Rank stability asks whether a district stays high over time. The Gi* test asks whether a district and its adjacent districts are jointly unusually high. Failing the false discovery rate threshold does not show that local clustering is absent. We therefore recommend describing the northeast (Habiganj, Netrakona, Sunamganj and Sylhet) and the southeast (Feni, Cumilla, and Noakhali) as persistently high areas, not hotspots.

Regression to the mean could affect the district classes, because a district classed high partly by chance will tend to read lower in the next round. Our resampling checks measure this noise but do not remove it. A set built from the 2009 survey located exposed people about as well, so the MICS rounds confirm the known map more than they redraw it. We chose the targeting rule during analysis, so its 2025 performance is not a prospective validation.

This concentration supports starting well testing in districts high in both earlier rounds, although it describes geographic coverage, not programme effectiveness or health impact. More than a quarter of exposed people lived outside these districts, and they were poorer. Geographic priority should add to open access to testing, not decide who is eligible. Testing should therefore continue elsewhere, on request and for every new well [31]. Myanmar shows a similar pattern under a 50 µg/L standard. National screening there found 8% of 123,962 wells above 50 µg/L but 71% above 10 µg/L, and the authors called for blanket screening focused on hotspot regions [38]. In Bangladesh the gap was narrower (14.4% against 8.6% of households in 2025), yet about two in five households above 10 ppb would still pass the 50 ppb standard.

Deep wells are often proposed as the safe source [39], but our comparison is cross-sectional. Where deep wells exist depends on place and on how community wells were allocated [40]. The ratio therefore cannot show that installing a deep well lowers arsenic, and local hydrogeology may confound it. The advantage also differed by division, so a deep well should not be assumed arsenic-safe without testing. Shallow wells low in arsenic were more often contaminated with faecal bacteria [41], and children drinking from them had more diarrhoea [42]. Yet studies in Matlab, a rural subdistrict of Chandpur, found 17% to 46% less childhood diarrhoea in households using deep tube wells [43,44]. Programmes that move households to safer wells therefore need to monitor microbial quality too.

Other studies suggest three steps to evaluate in the high districts. They are free testing of every new and untested well, durable labelling with education, and siting community wells by test results. In one upazila, more than 80% of wells installed after the national campaign were untested [31]. After testing, labelling, education and deep wells, 58% of participants with wells above 50 µg/L switched [39], against 27% after the national campaign alone [11]. The figures are from different sources, and so the gap is not an effect size. In Araihazar, only 29% of high-arsenic shallow wells were within 100 m of a community well [40]. Households need a safe source nearby to act on a test. Future MICS rounds should use a kit that performs better against laboratory results [18] and publish its laboratory cross-check. They should also report finer readings near 10 ppb and the month of each test.

This study used three nationally representative rounds, design-based estimates and the survey’s own laboratory check. It reproduced the published survey tables, which checks our computation but not the accuracy of the kits. The main limitation is that the later kits had no laboratory check, so the size of the fall rests on an untested assumption. The 2012-13 laboratory table has unpublished per-reading counts and partly counts from targeted samples, although wide changes to it did not remove the fall at 10 ppb. Three 2012-13 district values are doubtful. Replacing them strengthened the targeting result, but only as a sensitivity analysis. Fieldwork months differed between rounds, and the 2025 design tested two sources per cluster rather than one.

Wealth quintiles are relative within each round, and households reported well depth, in 2025 only. The maps describe sampled clusters at displaced coordinates. No health outcome or individual dose was measured. Source water may also differ from the water households store and drink, although the two agreed closely in 2019.

Arsenic contamination of household drinking-water sources in Bangladesh was lower in 2025 than in 2012-13. The size of the fall is uncertain without laboratory checks of the later kits. The fall did not redraw the map, and districts high in earlier rounds still held most exposed people. Testing should start there and continue elsewhere.

## Data availability

The survey data underlying this study are third-party data owned by UNICEF and cannot be shared by the authors. The datasets used were the MICS5 (2012-13), MICS6 (2019) and MICS7 (2025) household files. The MICS microdata are available to registered users who request them from the UNICEF MICS programme (https://mics.unicef.org), and the authors had no special access. The 2025 cluster global positioning system (GPS) file and the geospatial covariates are released by the MICS programme on request. District boundaries are open data from geoBoundaries (https://www.geoboundaries.org).

## Acknowledgments

We thank the UNICEF MICS programme and the Bangladesh Bureau of Statistics for the survey data.

## Supporting information

**S1 Fig.**
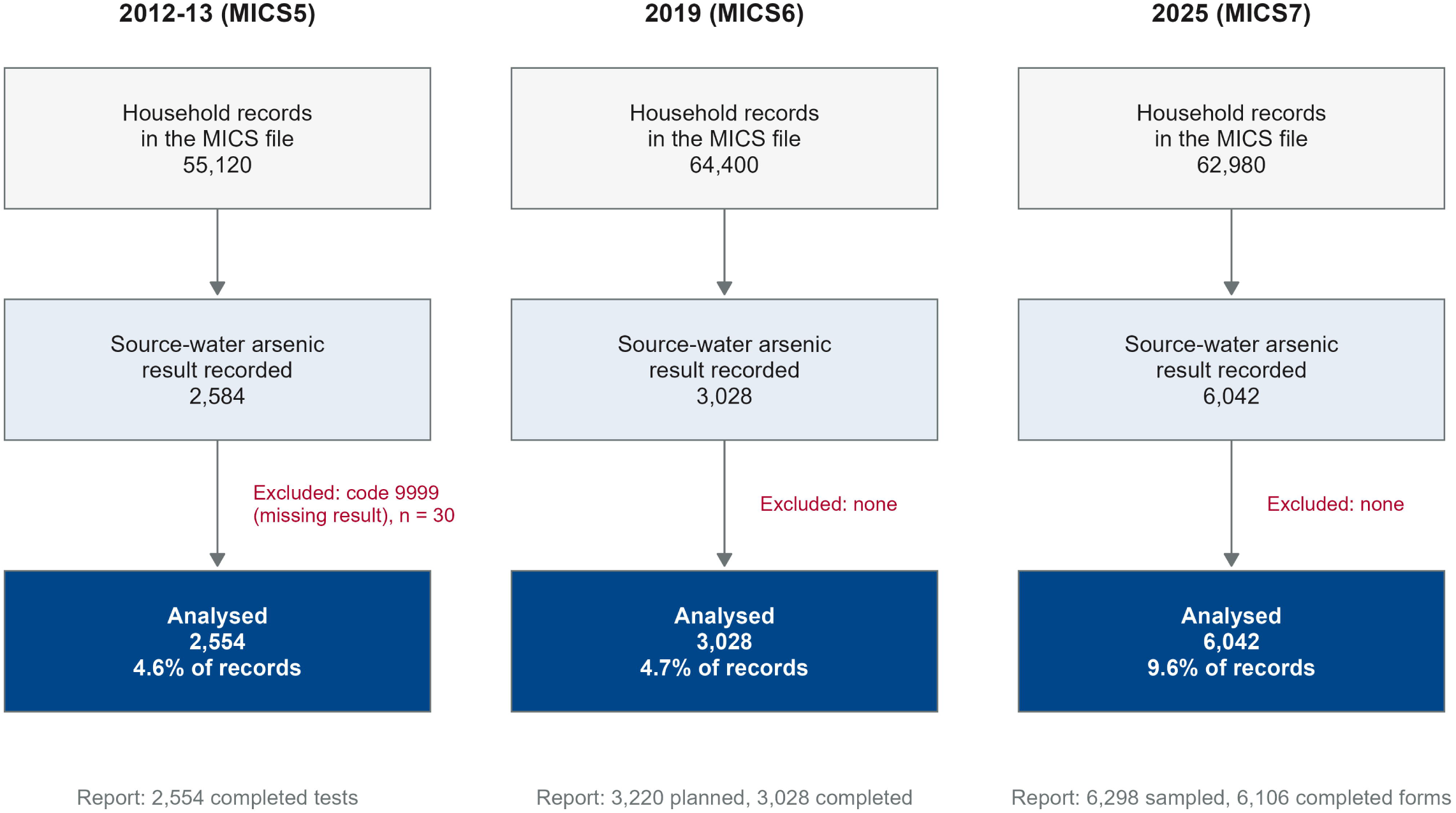
Participant flow by survey round.

**S2 Fig.**
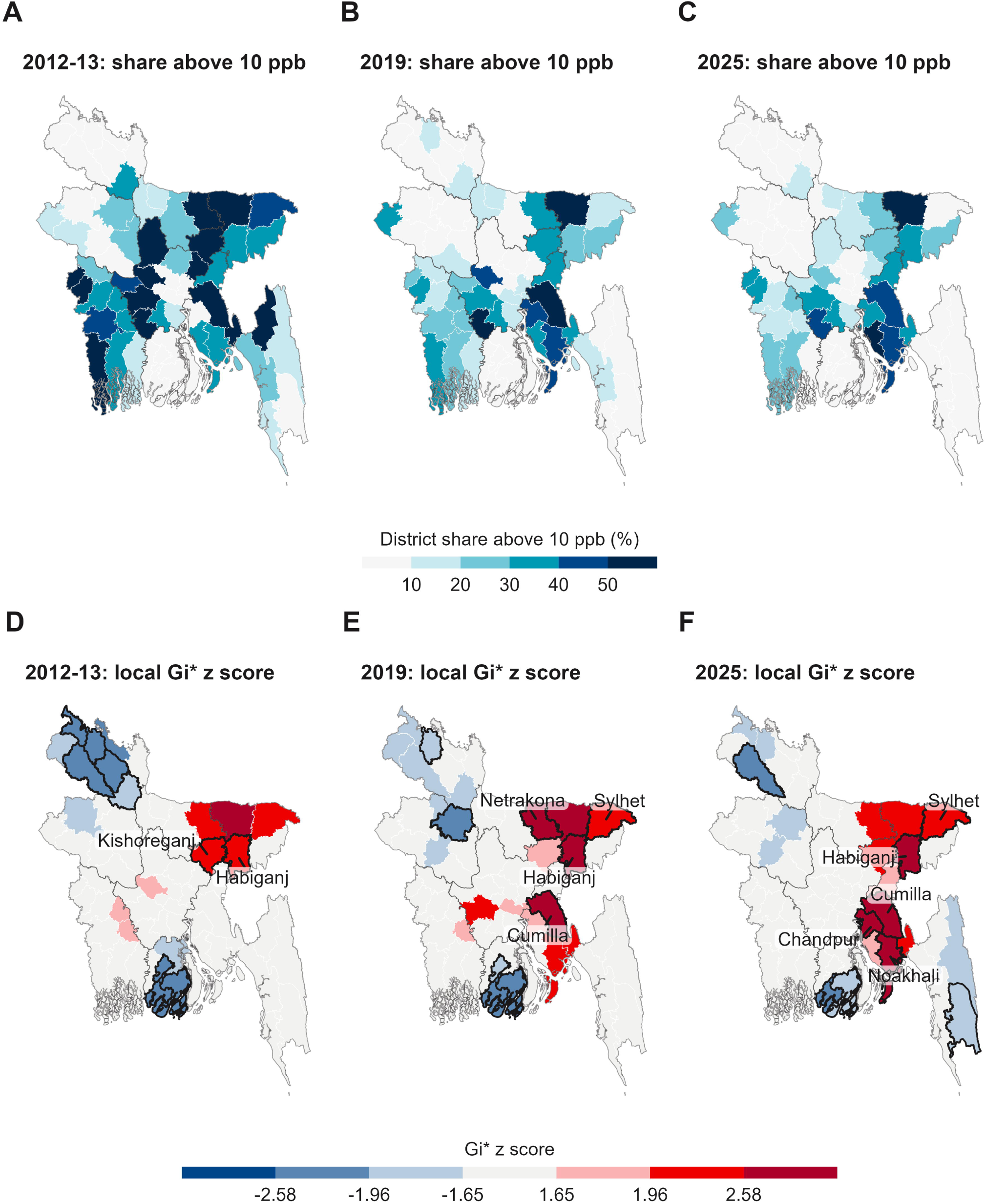
District arsenic above 10 ppb and local Getis-Ord Gi* by round.

**S2 Fig.**
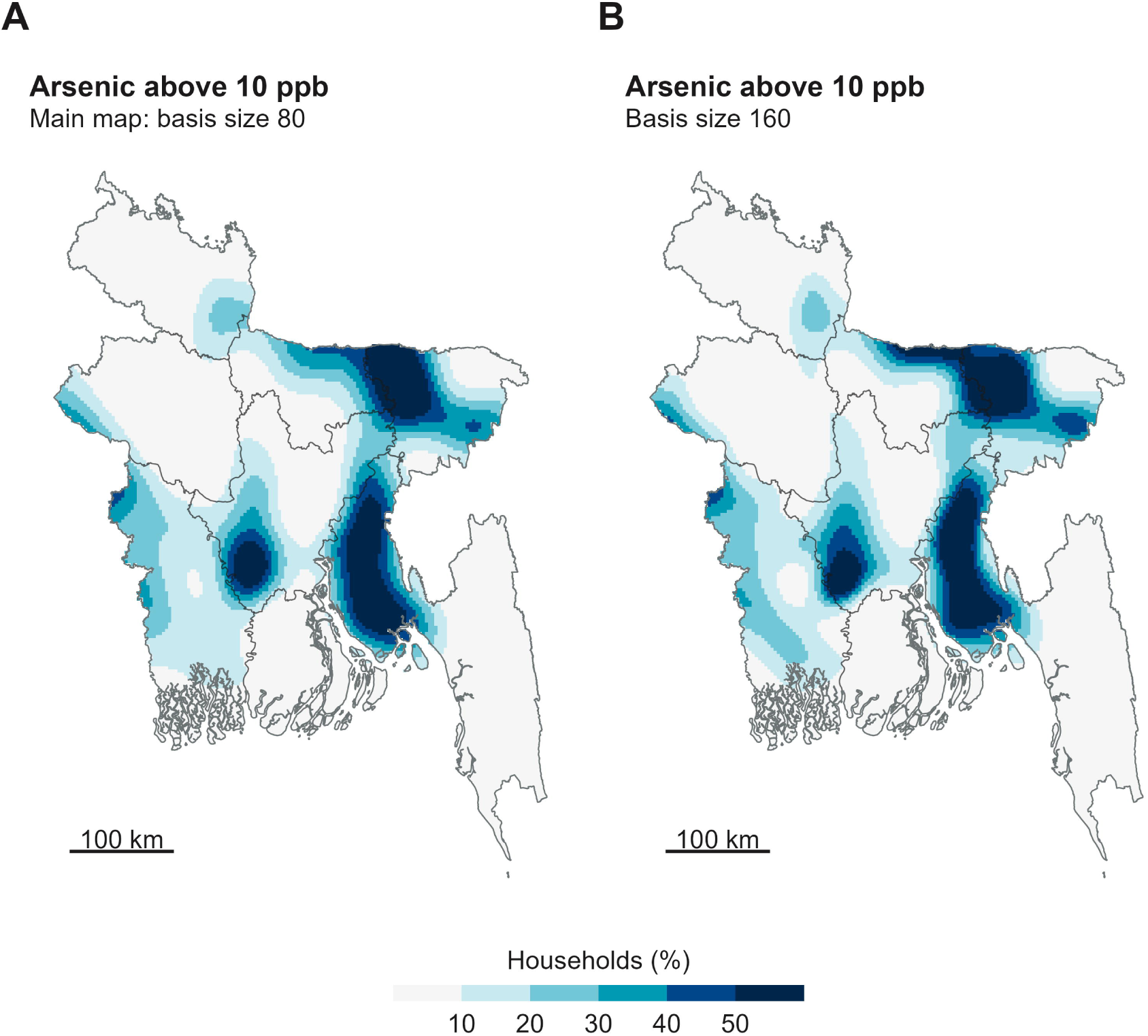
Sensitivity of the 2025 smoothed map to the spline basis size.

**S1 Table.** Kit readings and blank tests.

**S2 Table.** Laboratory cross-check of 2012-13 kit readings, with the weighted distribution of readings.

**S3 Table.** Kit and laboratory error scenarios.

**S4 Table.** Arsenic by main drinking-water source and round, with a split of the national change.

**S5 Table.** Robustness of the district targeting result.

**S6 Table.** Validation against the published MICS tables.

**S7 Table.** District prevalence above 10 ppb with 95% interval, 62 districts with at least 30 tests in every round.

**S8 Table.** Comparison with the district estimates of the 2009 national drinking water quality survey.

**S9 Table.** Households above 10 ppb by reported well depth and division, with adjusted ratios, 2025.

**S1 Text.** Additional methods and results.

**S1 Checklist.** STROBE checklist for cross-sectional studies.

